# Predict the next moves of COVID-19: reveal the temperate and tropical countries scenario

**DOI:** 10.1101/2020.04.04.20052928

**Authors:** Neaz A. Hasan, Mohammad Mahfujul Haque

## Abstract

The spread of COVID-19 engulfs almost all the countries and territories of the planet, and infections and fatality are increasing rapidly. The first epi-center of its’ massive spread was in Wuhan, Hubei province, China having a temperate weather, but the spread has got an unprecedented momentum in European temperate countries mainly in Italy and Spain (as of March 30, 2020). However, Malaysia and Singapore and the neighboring tropical countries of China got relatively low spread and fatality that created a research interest on whether there are potential impacts of weather condition on COVID-19 spread. Adopting the SIR (Susceptible Infected Removed) deviated model to predict potential cases and death in the coming days from COVID-19 was done using the secondary and official sources of data. This study shows that COVID-19 spread and fatality tend to be high across the world but compared to tropical countries, it is going to be incredibly high in the temperate countries having lower temperature (7-16°C) and humidity (80-90%) in last March. However, some literature predicted that this might not to be true, rather irrespective of weather conditions there might be a continuous spread and death. Moreover, a large number of asymptotic COVID-19 carrier in both temperate and tropical countries may re-outbreak in the coming winter. Therefore, a comprehensive global program with the leadership of WHO for testing of entire population of the world is required, which will be very useful for the individual states to take proper political action, social movement and medical services.

## Introduction

After the incidence of some mysterious SARS-like cluster pneumonia, cases with unknown aetiology on December 31, 2019 reported in Hubei province of Wuhan, China, novel coronavirus (2019-nCoV) was identified and characterized a few days later from that cases by the Chinese authorities (1,2). The disease is renamed as COVID-19 by World Health Organization (WHO) and virus name in transformed to severe acute respiratory syndrome coronavirus 2 (SARS-CoV- 2) by Coronavirus Study Group (CSG) of the International Committee on Virus Taxonomy, respectively on the same date 11 February, 2020 (3,4). Pointing over the 118,000 cases of illness by SARS-CoV-2 in over 110 countries WHO declared COVID-19 as pandemic on March 11, 2020 (5). Till date (March 30, 2020) the unstoppable virus is now emergent to 202 countries around the world and 2 international conveyances by grabbing 858,355 persons with the result of 42,309 death (6).

Wuhan, the center of coronavirus easing of its’ ‘City Closure’ situation after a lap of two months since 23 January, 2020 for successful attempts of the Chinese Government (7,8), when the other countries have newly been imposed the lockdown for uncertain period of time. The recent claim of China that the new COVID-19 cases essentially to zero has become possible for their quick action of test widely to isolate infected ones, stunted peoples’ unpopular movement except to get food and medical care, prohibit the leaving of own cities even homes of people and more importantly accepting the deep economy damage shut downed whole processing industry. The peak and quick recovery of the China made relax the rest of the world particularly the European countries. Moreover, Government of European countries’ emphasize attitude towards public opinion for being of affluent democracies and peoples’ free movement, easy travel and independent decision-making, which is now paying a price by counting 100,000 COVID-19 confirmed cases and 5,000 deaths with high fatality rate each day – expecting that the worst is yet to come (9).

By defying the geographical and political boundaries of western and temperate countries (like Italy, Spain, UK, USA etc.) the novel coronavirus has invaded towards tropical countries, excluding only a fewer than two dozen countries on the earth where there are no COVID-19 cases reported, so far. However, growth rate of conformed cases and fatalities are apparently slower in Asian tropical countries (like Malaysia, Singapore, Vietnam, Thailand etc.) than that of European temperate countries (10). This gap might be due to tropical countries with moderate economy are tended to be resource poor, not capable to do regular testing or there might be an impact of tropical weather to reduce the pandemic nature COVID-19, rather be a panic. A number of studies (11,12) argued that higher temperature and high relative humidity reduce the spreading capacity of new coronavirus to a greater extent, while there it would be popped again in 2020-2021 winter season (13). Researchers along with epidemiological experts also predicting COVID-19 cannot take advantage and bridgehead in the tropics than in the world’s temperate regions (14). A spatial analysis conducted by a research team from University of Maryland revealed that the transmission of COVID-19 lies mostly the regions across the equator where temperature ranges between 05-11°C with the combination of low specific (3-6 g/kg) and absolute humidity (4-7 g/m^3^) (15).

Depending on this complex context of COVID-19, this study is outlined to unpack the answers of the following research questions and formulate some recommendations to minimize the spread of COVID-19.

- What is the general trend of predictive cases and fatalities caused by Covid19 until end of April 2020 across the world?
- Is there any difference between the trends of the tropical and temperate countries?
- If there is a difference or otherwise, what are the recommendation to curve the spread of Covid19?

## Methods

Since the outbreak of coronavirus disease an exhaustive literature review related to the different aspects of COVID-19 and its’ relationships with different climatologic parameters and further forecasting was carried out. Published manuscripts, reports from different competent authorities, database and other documents on key areas of COVID-19 in different parts of the world were identified through searches in online journal sources and other available electronic sources.

## Model adopted

The proposed SIR (Susceptible Infected Removed) deviated model developed by Choi and Pak (16) during the outbreak of SARS on 2003, was adopted in this study to make a prediction about the suspected cases and death from being the onset of COVID-19. The COVID-19 transmission pattern from person-to-person, no pint source nature and negligible amount of immunity development by the exposure patient during the inaugural stage of COVID-19 pandemic is more likely to SARS epidemic. Therefore, the same model was adopted using the same four available parameters basic reproduction number (R_0_), case fatality rate (F), incubation period (i) and duration of disease (d) regarding COVID-19 revealed by other researchers.

## COVID-19 cases prediction

Basic reproductive number (R_0_) and incubation period (i) requires to play this model. Any disease become epidemic in any population of a geographical area by increasing the secondary cases from a single (typical) infection in that complete susceptible population is termed as basic reproduction number (17). And the incubation period is the time laps between initial infection and disease onset (18). The R_0_ will be changed by the increasing of i. Following the first incubation period, the total number of cases will be 1+ R_0_ for being the producing of R_0_ new cases from the single infectious case at the very beginning of the disease forming its’ epidemic nature. This will follow the quadrilateral nature in the subsequent periods of incubation and the total cases will be as 1 + (1 + R_0_) + (1 + R_0_ + R_0_^2^) + …………. + (1 + R_0_ + R_0_^2^ + ……… + R_0_^i^). If we consider C as the total number of predicted cases, then on t·i days the predicted number of incidents will be C_t_·_i_. Predicted total cases (C) of COVID-19 was calculated following the modified equation of Choi and Pak, (2003).

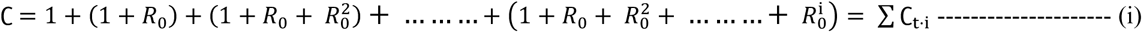

Equation (i) was tested by taking imaginary value of R_0_ =5 and i = 5 days that predict that on day 15 there will be 125 new cases with the resulting of total 156 cases (Table 1).

**Table 1.** Tryout the equation for predicting COVID-19 cases number

| Incubation period (t) | Day (t·i) | Predicted new cases ($C_{ti}$) | Predicted total cases (C) |
| --- | --- | --- | --- |
| 0 | 0 | 1* | 1 |
| 1 | 5 | 5 | 6 |
| 2 | 10 | 25 | 31 |
| 3 | 15 | 125 | 156 |

\* $R_0^0 = 5^0 = 1$

## COVID-19 death prediction

Like the case prediction of any epidemic disease, death prediction can also be done by a model using some parameters like basic reproductive number (R_0_), incubation period (i), duration of disease (d) and case fatality rate (F). Among these the first two is same of the case prediction model. Duration of disease (d) is the average time to diagnose people either cured or die. In more simple words maximum days to onset symptoms of the disease and if not disease diagnosed release the patient (19). The case fatality rate is the proportion of deaths within a defined population group (20). If D is considered as the predicted total deaths, the predicted deaths on t·i+d day can be calculated following the equation developed by Choi and Pak, (2003) during their prediction of SARS deaths.

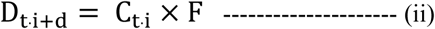

For predicting the total number of deaths, it is needed just to sum of all values get from the equation (ii). Therefore, the predicted total number of deaths is

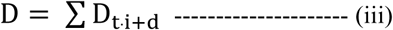

A trial was done for equation (ii) with the same value taken for equation (i) and taking the value for F = 20% and d = 14 days. The attempt reveals the prediction that first 0.2 persons will die from day 0 cases on day 14. Cases from day 5 will give 1 new death prediction with a total 1.2 persons’ death on day 19 (Table 2).

**Table 2.** Tryout the equation for predicting COVID-19 deaths

| Incubation period (t) | Day (t·i) | Predicted new cases ($C_{ti}$) | Predicted new deaths ($D_{ti+d}$) | Predicted total deaths (D) |
| --- | --- | --- | --- | --- |
| 0 | 0 | 1 | 0 | 0 |
| 1 | 5 | 3.28 | 0 | 0 |
| 2 | 10 | 10.76 | 0 | 0 |
|  | 14 (= t·i+d) | 0 | 0.2 | 0.2 |
| 3 | 15 | 35.29 | 0 | 0.2 |
|  | 19 | 0 | 1 | 1.2 |

## Adjust the values of different parameters from epidemiological studies

The adopted equations require different values from the recent epidemiological studies concerning to COVID-19 to get the predicted new cases and new death number, until the researchers invent a perfect vaccine to stop its’ devastating deadly nature. The prediction of total cases from equation (i) here we used R_0_ = 2.4 (average) (21) and i = 5 days (22). Recent epidemiological studies on COVID-19 (22–24) were harnessed to attain the i value. From a number of other studies (25–27) several values of R_0_ were attained and trialed to check the perfect combination of i and R_0_. The perfect combination was determined by the closest match between the observed total number of COVID-19 cases reported by World Health Organization (WHO) and the attained total number of COVID-19 cases from equation (i). Besides these two values for equation (ii) and (iii) another two values namely duration of disease (d) and case fatality rate (F) were required. The disease duration for COVID-19 has been set 14 days by WHO (28) used in this study. This means the case from day 0 will be recovered or dead on day 14. Similarly, from second incubation period i.e. patient from day 5 recovered or dead on day 19 and so on. Case fatality rate (F) values of COVID-19 is differed in different studies (29–31), however to avoid the regional confinement, here in this study WHO reported COVID-19 case fatality rate (F) 3.4% (3) was used. This pointed that the death from the cases day 0 is 0.034 (1×3.4%) on day 14, as opposed to be recovered 0.966 people. The values of R_0_ and i selected in the was well adjusted with d and F values selected from WHOs’ report.

In case of determining the prediction for sub-tropical and tropical countries the incubation period (i) and duration of disease (d) values kept same, as these values do not vary by geographical variation for any kind of infectious disease (32). For basic reproduction number (R_0_) and fatality rate (F) the values revealed by different regional researchers were collected, checked with the i and d value for adjustment and finalized (Table 3).

**Table 3.** Adjusted basic reproduction number and fatality rate for sub-tropical and tropical countries

| Region | Countries | Basic reproduction number ($R_0$) | Fatality rate (F) |
| --- | --- | --- | --- |
| Sub-tropical | Italy | 3.001 | 11.39% |
|  |  | (51) | (52) |
|  | Spain | 3.001* | 8.67% |
|  |  |  | (52) |
| Tropical | Malaysia | 1.7 | 1.55% |
|  |  | (53) | (52) |
|  | Singapore | 1.1 | 0.3% |
|  |  | (54) | (29) |

\*For sharing the same tropical region and same trend is observed from WHO and Johns Hopkins University daily report the same value from Italian researchers was used for unavailable data of Spain from any trustworthy source.

## Data source and data analysis

It was required two different types of data. Since for the prediction of daily new cases and deaths from COVID-19 the data from the official website of WHO (https://www.who.int/emergencies/diseases/novel-coronavirus-2019) and Center for Systems Science and Engineering, Johns Hopkins University, USA (https://www.arcgis.com/apps/opsdashboard/index.html#/bda7594740fd40299423467b48e9ecf6) were used. The weather data of temperate and tropical countries were gathered from authentic website of Weather Atlas (https://www.weather-atlas.com). All the collected data was imputed, analyzed and generated results using Excel (Microsoft Corporation).

## Results

### Predicted cases and deaths of the world

Result of probable cases and deaths for being the onset of COVID-19 was attained based on the ongoing trend of the deadly virus SARS-CoV-2 in all over the world. The predicted result revealed 326.89 cases and 0.79 deaths (day 30; January 30, 2020); 62605.6 cases and 368.41 deaths (day 60; February 29, 2020); 11964251.42 cases and 70621.17 deaths (day 90; March 30, 2020) and 2286404188 cases and 13496134.68 deaths (day 120; April 29, 2020) in each quartile of 120 days prediction. The predicted cases and death results were very much resembled with the observed data from March 14, 2020 and onwards (Table 4).

**Table 4.**
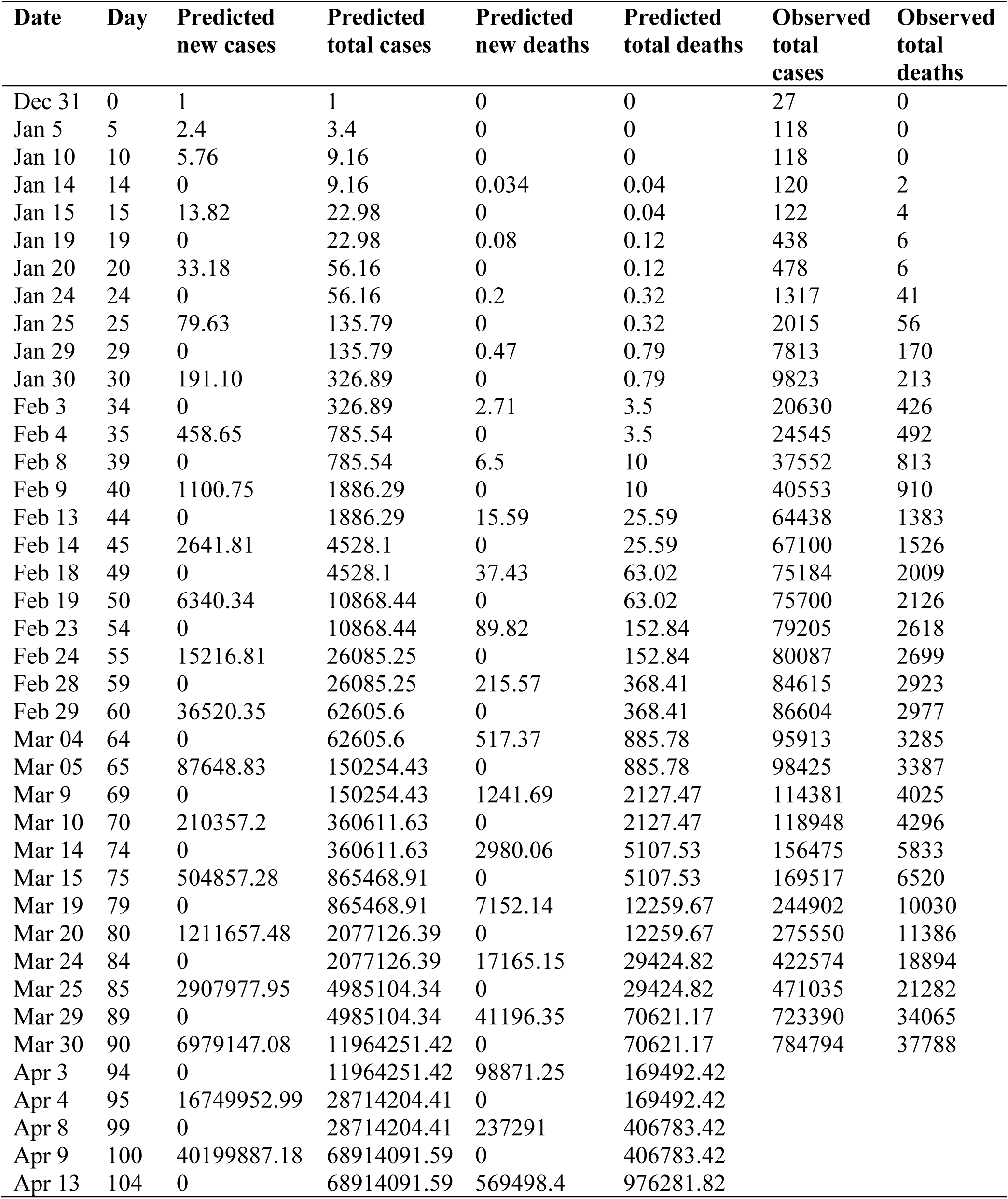
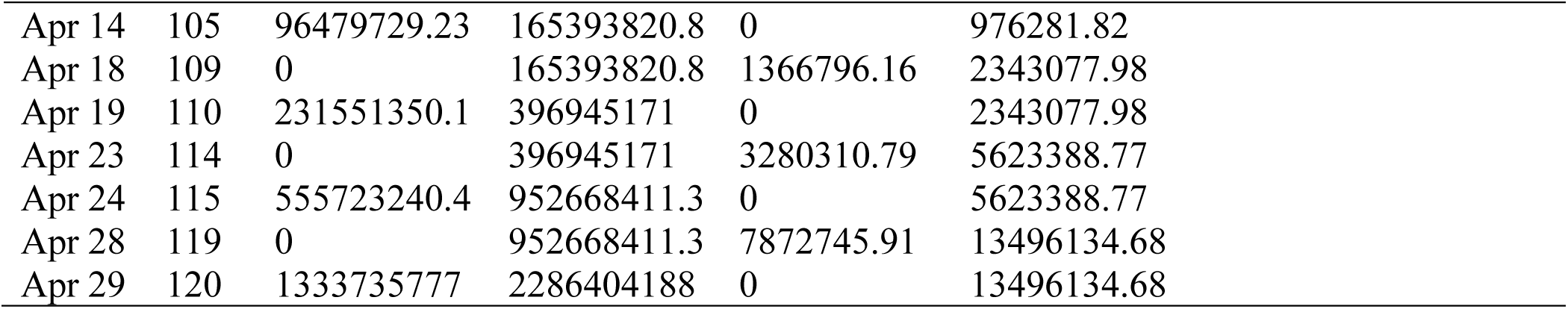
Probable predicted cases and deaths of COVID-19 after its’ first outbreak (December 31, 2019) in Wuhan, China based on the trend till February 29, 2020 and exposed number as of March 30, 2020

| **Date** | **Day** | **Predicted**<br>**new cases** | **Predicted**<br>**total cases** | **Predicted**<br>**new deaths** | **Predicted**<br>**total deaths** | **Observed**<br>**total**<br>**cases** | **Observed**<br>**total**<br>**deaths** |
| --- | --- | --- | --- | --- | --- | --- | --- |
| Dec 31 | 0 | 1 | 1 | 0 | 0 | 27 | 0 |
| Jan 5 | 5 | 2.4 | 3.4 | 0 | 0 | 118 | 0 |
| Jan 10 | 10 | 5.76 | 9.16 | 0 | 0 | 118 | 0 |
| Jan 14 | 14 | 0 | 9.16 | 0.034 | 0.04 | 120 | 2 |
| Jan 15 | 15 | 13.82 | 22.98 | 0 | 0.04 | 122 | 4 |
| Jan 19 | 19 | 0 | 22.98 | 0.08 | 0.12 | 438 | 6 |
| Jan 20 | 20 | 33.18 | 56.16 | 0 | 0.12 | 478 | 6 |
| Jan 24 | 24 | 0 | 56.16 | 0.2 | 0.32 | 1317 | 41 |
| Jan 25 | 25 | 79.63 | 135.79 | 0 | 0.32 | 2015 | 56 |
| Jan 29 | 29 | 0 | 135.79 | 0.47 | 0.79 | 7813 | 170 |
| Jan 30 | 30 | 191.10 | 326.89 | 0 | 0.79 | 9823 | 213 |
| Feb 3 | 34 | 0 | 326.89 | 2.71 | 3.5 | 20630 | 426 |
| Feb 4 | 35 | 458.65 | 785.54 | 0 | 3.5 | 24545 | 492 |
| Feb 8 | 39 | 0 | 785.54 | 6.5 | 10 | 37552 | 813 |
| Feb 9 | 40 | 1100.75 | 1886.29 | 0 | 10 | 40553 | 910 |
| Feb 13 | 44 | 0 | 1886.29 | 15.59 | 25.59 | 64438 | 1383 |
| Feb 14 | 45 | 2641.81 | 4528.1 | 0 | 25.59 | 67100 | 1526 |
| Feb 18 | 49 | 0 | 4528.1 | 37.43 | 63.02 | 75184 | 2009 |
| Feb 19 | 50 | 6340.34 | 10868.44 | 0 | 63.02 | 75700 | 2126 |
| Feb 23 | 54 | 0 | 10868.44 | 89.82 | 152.84 | 79205 | 2618 |
| Feb 24 | 55 | 15216.81 | 26085.25 | 0 | 152.84 | 80087 | 2699 |
| Feb 28 | 59 | 0 | 26085.25 | 215.57 | 368.41 | 84615 | 2923 |
| Feb 29 | 60 | 36520.35 | 62605.6 | 0 | 368.41 | 86604 | 2977 |
| Mar 04 | 64 | 0 | 62605.6 | 517.37 | 885.78 | 95913 | 3285 |
| Mar 05 | 65 | 87648.83 | 150254.43 | 0 | 885.78 | 98425 | 3387 |
| Mar 9 | 69 | 0 | 150254.43 | 1241.69 | 2127.47 | 114381 | 4025 |
| Mar 10 | 70 | 210357.2 | 360611.63 | 0 | 2127.47 | 118948 | 4296 |
| Mar 14 | 74 | 0 | 360611.63 | 2980.06 | 5107.53 | 156475 | 5833 |
| Mar 15 | 75 | 504857.28 | 865468.91 | 0 | 5107.53 | 169517 | 6520 |
| Mar 19 | 79 | 0 | 865468.91 | 7152.14 | 12259.67 | 244902 | 10030 |
| Mar 20 | 80 | 1211657.48 | 2077126.39 | 0 | 12259.67 | 275550 | 11386 |
| Mar 24 | 84 | 0 | 2077126.39 | 17165.15 | 29424.82 | 422574 | 18894 |
| Mar 25 | 85 | 2907977.95 | 4985104.34 | 0 | 29424.82 | 471035 | 21282 |
| Mar 29 | 89 | 0 | 4985104.34 | 41196.35 | 70621.17 | 723390 | 34065 |
| Mar 30 | 90 | 6979147.08 | 11964251.42 | 0 | 70621.17 | 784794 | 37788 |
| Apr 3 | 94 | 0 | 11964251.42 | 98871.25 | 169492.42 |  |  |
| Apr 4 | 95 | 16749952.99 | 28714204.41 | 0 | 169492.42 |  |  |
| Apr 8 | 99 | 0 | 28714204.41 | 237291 | 406783.42 |  |  |
| Apr 9 | 100 | 40199887.18 | 68914091.59 | 0 | 406783.42 |  |  |
| Apr 13 | 104 | 0 | 68914091.59 | 569498.4 | 976281.82 |  |  |

|  |  |  |  |  |  |
| --- | --- | --- | --- | --- | --- |
| Apr 14 | 105 | 96479729.23 | 165393820.8 | 0 | 976281.82 |
| Apr 18 | 109 | 0 | 165393820.8 | 1366796.16 | 2343077.98 |
| Apr 19 | 110 | 231551350.1 | 396945171 | 0 | 2343077.98 |
| Apr 23 | 114 | 0 | 396945171 | 3280310.79 | 5623388.77 |
| Apr 24 | 115 | 555723240.4 | 952668411.3 | 0 | 5623388.77 |
| Apr 28 | 119 | 0 | 952668411.3 | 7872745.91 | 13496134.68 |
| Apr 29 | 120 | 1333735777 | 2286404188 | 0 | 13496134.68 |

### COVID-19 scenario of the tropical countries and sub-tropical countries

During the writeup of this study Italy and Spain are the two most vulnerable sub-tropical countries selected for this study to tale the COVID-19 incidental difference between sub-tropical and tropical countries. After the exact one-month laps of the first incidence of COVID-19 the deadly virus in Wuhan, China; the first reported case in for both Italy and Spain was on January 31, 2020. The result is presented of 90 days, while there is arrayed comparative results of predicted and observed for two months and another one month only predicted results. Predicted cases of both Italy and Spain was same as we used the same basic reproduction value for being of the countries from the same temperate and geographical position. However, the predicted total death varies across the countries as the fatality rate was not same (Table 5 and 6).

**Table 5.** Probable predicted cases and deaths of COVID-19 after its’ first outbreak (January 31, 2020) in Italy based on the trend and exposed number till March 31, 2020

| **Date** | **Day** | **Predicted**<br>**new cases** | **Predicted**<br>**total cases** | **Predicted**<br>**new deaths** | **Predicted**<br>**total deaths** | **Observed**<br>**total**<br>**cases** | **Observed**<br>**total**<br>**deaths** |
| --- | --- | --- | --- | --- | --- | --- | --- |
| Jan 31 | 0 | 1 | 1 | 0 | 0 | 2 | 0 |
| Feb 5 | 5 | 3 | 4 | 0 | 0 | 2 | 0 |
| Feb 10 | 10 | 9.01 | 13.01 | 0 | 0 | 3 | 0 |
| Feb 14 | 14 | 0 | 13.01 | 0.11 | 0.11 | 3 | 0 |
| Feb 15 | 15 | 27.02 | 40.03 | 0 | 0.11 | 3 | 0 |
| Feb 19 | 19 | 0 | 40.03 | 0.34 | 0.45 | 3 | 0 |
| Feb 20 | 20 | 81.11 | 121.14 | 0 | 0.45 | 3 | 0 |
| Feb 24 | 24 | 0 | 121.14 | 1.03 | 1.48 | 229 | 7 |
| Feb 25 | 25 | 243.40 | 364.54 | 0 | 1.48 | 322 | 11 |
| Feb 29 | 29 | 0 | 364.54 | 3.08 | 4.56 | 1128 | 29 |
| Mar 1 | 30 | 730.46 | 1095 | 0 | 4.56 | 1701 | 41 |
| Mar 5 | 34 | 0 | 1095 | 27.75 | 32.31 | 3858 | 148 |
| Mar 6 | 35 | 2192.11 | 3287.11 | 0 | 32.31 | 4636 | 197 |
| Mar 10 | 39 | 0 | 3287.11 | 83.27 | 115.58 | 10149 | 631 |
| Mar 11 | 40 | 6578.52 | 9865.63 | 0 | 115.58 | 12462 | 827 |
| Mar 15 | 44 | 0 | 9865.63 | 249.9 | 365.48 | 24747 | 1809 |
| Mar 16 | 45 | 19742.12 | 29607.75 | 0 | 365.48 | 27980 | 2158 |
| Mar 20 | 49 | 0 | 29607.75 | 749.95 | 1115.43 | 47021 | 4032 |
| Mar 21 | 50 | 59246.13 | 88853.88 | 0 | 1115.43 | 53578 | 4825 |
| Mar 25 | 54 | 0 | 88853.88 | 2250.60 | 3366.03 | 74386 | 7503 |
| Mar 26 | 55 | 177797.62 | 266651.5 | 0 | 3366.03 | 80589 | 8215 |
| Mar 30 | 59 | 0 | 266651.5 | 6754.06 | 10120.09 | 101739 | 11591 |
| Mar 31 | 60 | 533570.67 | 800222.17 | 0 | 10120.09 | 105792 | 12428 |
| Apr 4 | 64 | 0 | 800222.17 | 20268.93 | 30389.02 |  |  |
| Apr 5 | 65 | 1601245.57 | 2401467.74 | 0 | 30389.02 |  |  |
| Apr 9 | 69 | 0 | 2401467.74 | 60827.06 | 91216.08 |  |  |
| Apr 10 | 70 | 4805337.94 | 7206805.68 | 0 | 91216.08 |  |  |
| Apr 14 | 74 | 0 | 7206805.68 | 182541.99 | 273758.07 |  |  |
| Apr 15 | 75 | 14420819.18 | 21627624.86 | 0 | 273758.07 |  |  |
| Apr 19 | 79 | 0 | 21627624.86 | 547808.53 | 821566.6 |  |  |
| Apr 20 | 80 | 43276878.34 | 64904503.2 | 0 | 821566.6 |  |  |
| Apr 24 | 84 | 0 | 64904503.2 | 1643973.38 | 2465539.98 |  |  |
| Apr 25 | 85 | 129873912 | 194778415.2 | 0 | 2465539.98 |  |  |
| Apr 29 | 89 | 0 | 194778415.2 | 4933564.13 | 7399104.11 |  |  |
| Apr 30 | 90 | 389751609.8 | 584530025 | 0 | 7399104.11 |  |  |

**Table 6.** Probable predicted cases and deaths of COVID-19 after its’ first outbreak (January 31, 2020) in Spain based on the trend and exposed number till March 31, 2020

| **Date** | **Day** | **Predicted**<br>**new cases** | **Predicted**<br>**total cases** | **Predicted**<br>**new deaths** | **Predicted**<br>**total deaths** | **Observed**<br>**total**<br>**cases** | **Observed**<br>**total**<br>**deaths** |
| --- | --- | --- | --- | --- | --- | --- | --- |
| Jan 31 | 0 | 1 | 1 | 0 | 0 | 1 | 0 |
| Feb 5 | 5 | 3 | 4 | 0 | 0 | 1 | 0 |
| Feb 10 | 10 | 9.01 | 13.01 | 0 | 0 | 1 | 0 |
| Feb 14 | 14 | 0 | 13.01 | 0.09 | 0.09 | 2 | 0 |
| Feb 15 | 15 | 27.02 | 40.03 | 0 | 0.09 | 2 | 0 |
| Feb 19 | 19 | 0 | 40.03 | 0.27009 | 0.36 | 2 | 0 |
| Feb 20 | 20 | 81.11 | 121.14 | 0 | 0.36 | 2 | 0 |
| Feb 24 | 24 | 0 | 121.14 | 0.81054009 | 1.17 | 2 | 0 |
| Feb 25 | 25 | 243.40 | 364.54 | 0 | 1.17 | 6 | 0 |
| Feb 29 | 29 | 0 | 364.54 | 2.43243081 | 3.6 | 45 | 0 |
| Mar 1 | 30 | 730.46 | 1095 | 0 | 3.6 | 84 | 0 |
| Mar 5 | 34 | 0 | 1095 | 21.9 | 25.5 | 259 | 3 |
| Mar 6 | 35 | 2192.11 | 3287.11 | 0 | 25.5 | 400 | 8 |
| Mar 10 | 39 | 0 | 3287.11 | 65.75 | 91.25 | 1695 | 36 |
| Mar 11 | 40 | 6578.52 | 9865.63 | 0 | 91.25 | 2277 | 55 |
| Mar 15 | 44 | 0 | 9865.63 | 197.29 | 288.54 | 7988 | 294 |
| Mar 16 | 45 | 19742.12 | 29607.75 | 0 | 288.54 | 9942 | 342 |
| Mar 20 | 49 | 0 | 29607.75 | 592.06 | 880.6 | 21571 | 1093 |
| Mar 21 | 50 | 59246.13 | 88853.88 | 0 | 880.6 | 25496 | 1381 |
| Mar 25 | 54 | 0 | 88853.88 | 1776.79 | 2657.39 | 49515 | 3647 |
| Mar 26 | 55 | 177797.62 | 266651.5 | 0 | 2657.39 | 57786 | 4365 |
| Mar 30 | 59 | 0 | 266651.5 | 5332.15 | 7989.54 | 87956 | 7716 |
| Mar 31 | 60 | 533570.67 | 800222.17 | 0 | 7989.54 | 95923 | 8464 |
| Apr 4 | 64 | 0 | 800222.17 | 16001.79 | 23991.33 |  |  |
| Apr 5 | 65 | 1601245.57 | 2401467.74 | 0 | 23991.33 |  |  |
| Apr 9 | 69 | 0 | 2401467.74 | 48021.36 | 72012.69 |  |  |
| Apr 10 | 70 | 4805337.94 | 7206805.68 | 0 | 72012.69 |  |  |
| Apr 14 | 74 | 0 | 7206805.68 | 144112.1 | 216124.79 |  |  |
| Apr 15 | 75 | 14420819.18 | 21627624.86 | 0 | 216124.79 |  |  |
| Apr 19 | 79 | 0 | 21627624.86 | 432480.42 | 648605.21 |  |  |
| Apr 20 | 80 | 43276878.34 | 64904503.2 | 0 | 648605.21 |  |  |
| Apr 24 | 84 | 0 | 64904503.2 | 1297873.72 | 1946478.93 |  |  |
| Apr 25 | 85 | 129873912 | 194778415.2 | 0 | 1946478.93 |  |  |
| Apr 29 | 89 | 0 | 194778415.2 | 3894919.05 | 5841397.98 |  |  |
| Apr 30 | 90 | 389751609.8 | 584530025 | 0 | 5841397.98 |  |  |

Malaysia and Singapore are selected as representing countries from tropical listed countries affected by COVID-19. The estimated result for Malaysia was 57.17 cases and 0.21 deaths (day 30; February 24, 2020); 1413.5 cases and 9.59 deaths (day 60; March 25, 2020) against the observed result of 22 cases with 0 deaths and 1796 cases with 20 deaths on the same date of 30 days interval. The predicted result for next 30 days interval (day 90, April 24, 2020) is 34151.76 cases with 236.15 deaths (Table 7). The computed probable result for another tropical country Singapore was 9.48 cases and 0.014 deaths (day 30; February 22, 2020); 24.52 cases and 0.051 deaths (day 60; March 23, 2020) when 85 cases with 0 deaths and 509 cases with 2 deaths were observed on the same 30 days interval date. And the prediction for the day 90 (April 22, 2020) is 51.15 cases with negligible number of death (0.11) (Table 8).

**Table 7.** Probable predicted cases and deaths of COVID-19 after its’ first outbreak (January 31, 2020) in Malaysia based on the trend and exposed number till March 30, 2020

| **Date** | **Day** | **Predicted**<br>**new cases** | **Predicted**<br>**total cases** | **Predicted**<br>**new deaths** | **Predicted**<br>**total deaths** | **Observed**<br>**total**<br>**cases** | **Observed**<br>**total**<br>**deaths** |
| --- | --- | --- | --- | --- | --- | --- | --- |
| Jan 25 | 0 | 1 | 1 | 0 | 0 | 3 | 0 |
| Jan 30 | 5 | 1.7 | 2.7 | 0 | 0 | 8 | 0 |
| Feb 4 | 10 | 2.89 | 5.59 | 0 | 0 | 10 | 0 |
| Feb 8 | 14 | 0 | 5.59 | 0.02 | 0.02 | 16 | 0 |
| Feb 9 | 15 | 4.91 | 10.5 | 0 | 0.02 | 17 | 0 |
| Feb 13 | 19 | 0 | 10.5 | 0.03 | 0.05 | 19 | 0 |
| Feb 14 | 20 | 8.35 | 18.85 | 0 | 0.05 | 19 | 0 |
| Feb 18 | 24 | 0 | 18.85 | 0.06 | 0.11 | 22 | 0 |
| Feb 19 | 25 | 14.2 | 33.05 | 0 | 0.11 | 22 | 0 |
| Feb 23 | 29 | 0 | 33.05 | 0.1 | 0.21 | 22 | 0 |
| Feb 24 | 30 | 24.14 | 57.19 | 0 | 0.21 | 22 | 0 |
| Feb 28 | 34 | 0 | 57.19 | 0.28 | 0.49 | 25 | 0 |
| Feb 29 | 35 | 41.03 | 98.22 | 0 | 0.49 | 29 | 0 |
| Mar 04 | 39 | 0 | 98.22 | 0.48 | 0.97 | 50 | 0 |
| Mar 05 | 40 | 69.76 | 167.98 | 0 | 0.97 | 55 | 0 |
| Mar 9 | 44 | 0 | 167.98 | 0.82 | 1.79 | 117 | 0 |
| Mar 10 | 45 | 118.59 | 286.57 | 0 | 1.79 | 129 | 0 |
| Mar 14 | 49 | 0 | 286.57 | 1.4 | 3.19 | 238 | 0 |
| Mar 15 | 50 | 201.59 | 488.16 | 0 | 3.19 | 428 | 0 |
| Mar 19 | 54 | 0 | 488.16 | 2.37 | 5.56 | 900 | 3 |
| Mar 20 | 55 | 342.72 | 830.88 | 0 | 5.56 | 1030 | 4 |
| Mar 24 | 59 | 0 | 830.88 | 4.03 | 9.59 | 1624 | 16 |
| Mar 25 | 60 | 582.62 | 1413.5 | 0 | 9.59 | 1796 | 20 |
| Mar 29 | 64 | 0 | 1413.5 | 6.86 | 16.45 | 2470 | 35 |
| Mar 30 | 65 | 990.46 | 2403.96 | 0 | 16.45 | 2626 | 37 |
| Apr 3 | 69 | 0 | 2403.96 | 11.65 | 28.1 |  |  |
| Apr 4 | 70 | 1683.78 | 4087.74 | 0 | 28.1 |  |  |
| Apr 8 | 74 | 0 | 4087.74 | 19.81 | 47.91 |  |  |
| Apr 9 | 75 | 2862.43 | 6950.17 | 0 | 47.91 |  |  |
| Apr 13 | 79 | 0 | 6950.17 | 33.67 | 81.58 |  |  |
| Apr 14 | 80 | 4866.11 | 11816.28 | 0 | 81.58 |  |  |
| Apr 18 | 84 | 0 | 11816.28 | 57.25 | 138.83 |  |  |
| Apr 19 | 85 | 8272.4 | 20088.68 | 0 | 138.83 |  |  |
| Apr 23 | 89 | 0 | 20088.68 | 97.32 | 236.15 |  |  |
| Apr 24 | 90 | 14063.08 | 34151.76 | 0 | 236.15 |  |  |

**Table 8.** Probable predicted cases and deaths of COVID-19 after its’ first outbreak (January 31, 2020) in Singapore based on the trend and exposed number till March 31, 2020

| **Date** | **Day** | **Predicted**<br>**new cases** | **Predicted**<br>**total cases** | **Predicted**<br>**new deaths** | **Predicted**<br>**total deaths** | **Observed**<br>**total**<br>**cases** | **Observed**<br>**total**<br>**deaths** |
| --- | --- | --- | --- | --- | --- | --- | --- |
| Jan 23 | 0 | 1 | 1 | 0 | 0 | 1 | 0 |
| Jan 28 | 5 | 1.1 | 2.1 | 0 | 0 | 7 | 0 |
| Feb 2 | 10 | 1.21 | 3.31 | 0 | 0 | 18 | 0 |
| Feb 6 | 14 | 0 | 3.31 | 0.003 | 0.003 | 28 | 0 |
| Feb 7 | 15 | 1.331 | 4.64 | 0 | 0.003 | 30 | 0 |
| Feb 11 | 19 | 0 | 4.64 | 0.003 | 0.006 | 47 | 0 |
| Feb 12 | 20 | 1.46 | 6.1 | 0 | 0.006 | 50 | 0 |
| Feb 16 | 24 | 0 | 6.1 | 0.004 | 0.01 | 75 | 0 |
| Feb 17 | 25 | 1.61 | 7.71 | 0 | 0.01 | 77 | 0 |
| Feb 21 | 29 | 0 | 7.71 | 0.004 | 0.014 | 85 | 0 |
| Feb 22 | 30 | 1.77 | 9.48 | 0 | 0.014 | 85 | 0 |
| Feb 26 | 34 | 0 | 9.48 | 0.005 | 0.019 | 93 | 0 |
| Feb 27 | 35 | 1.95 | 11.43 | 0 | 0.019 | 93 | 0 |
| Mar 2 | 39 | 0 | 11.43 | 0.005 | 0.024 | 108 | 0 |
| Mar 3 | 40 | 2.14 | 13.57 | 0 | 0.024 | 110 | 0 |
| Mar 7 | 44 | 0 | 13.57 | 0.006 | 0.03 | 138 | 0 |
| Mar 8 | 45 | 2.36 | 15.93 | 0 | 0.03 | 150 | 0 |
| Mar 12 | 49 | 0 | 15.93 | 0.006 | 0.036 | 178 | 0 |
| Mar 13 | 50 | 2.6 | 18.53 | 0 | 0.036 | 200 | 0 |
| Mar 17 | 54 | 0 | 18.53 | 0.007 | 0.043 | 266 | 0 |
| Mar 18 | 55 | 2.85 | 21.38 | 0 | 0.043 | 313 | 0 |
| Mar 22 | 59 | 0 | 21.38 | 0.008 | 0.051 | 455 | 2 |
| Mar 23 | 60 | 3.14 | 24.52 | 0 | 0.051 | 509 | 2 |
| Mar 27 | 64 | 0 | 24.52 | 0.009 | 0.06 | 732 | 2 |
| Mar 28 | 65 | 3.45 | 27.97 | 0 | 0.06 | 802 | 2 |
| Apr 1 | 69 | 0 | 27.97 | 0.009 | 0.069 |  |  |
| Apr 2 | 70 | 3.8 | 31.77 | 0 | 0.069 |  |  |
| Apr 6 | 74 | 0 | 31.77 | 0.01 | 0.079 |  |  |
| Apr 7 | 75 | 4.17 | 35.94 | 0 | 0.079 |  |  |
| Apr 11 | 79 | 0 | 35.94 | 0.011 | 0.09 |  |  |
| Apr 12 | 80 | 4.59 | 40.55 | 0 | 0.09 |  |  |
| Apr 16 | 84 | 0 | 40.55 | 0.01 | 0.1 |  |  |
| Apr 17 | 85 | 5.05 | 45.59 | 0 | 0.1 |  |  |
| Apr 21 | 89 | 0 | 45.59 | 0.01 | 0.11 |  |  |
| Apr 22 | 90 | 5.56 | 51.15 | 0 | 0.11 |  |  |

## Discussion

The number of predicted cases and death was very much resembled after a certain time interval with that of the actual number of cases and death for both global and regional population, however it has some unexpected difference. Actual forecasting like “How many people will be affected onset by the following days” or “The outbreak going to be peak or not in the coming month?” of an epidemic disease from a transmission dynamics based model is quite tough (33). This is because the transmission dynamics based model is developed foothold on the official clinical surveillance data, which comes through a bureaucratic reporting chain of public health officials and government employees (34) who have to maintain privacy concern owing to geographical resolutions. Moreover, human (host) social behavior (35), potential changes in the pathogen (36) and host-pathogen interaction (37) dominates the epidemic models to a greater extent, while these are continuously change within an epidemic period. In case of COVID-19 the prediction from this study can be rippled for being down warding the basic reproduction value, which is already slowed from 2.9 to 2.4 (26) and also of the fatality rate which must be slow downed on the coming days for taking effective measure by the affected countries.

Using the existing value of different parameters and incorporated the values in the epidemic model we revealed an estimation of global total cases (2286404188) and death numbers (13496134.68) on April 29, 2020. By contrast with the SARS-2003 epidemic and MARS-2012 outbreak, this estimation of COVID-19 pandemic got some furious figure for its’ haughty R_0_ value (median 2.4) from the very early devastating nature of the virus, where SARS-2003 and MARS-2012 R_0_ values were 0.19–1.08 (38) and 0.29–0.80 (39), respectively. Moreover, this unbelievable number can be a true figure if this virus continues its’ sabotage, and we observe the present situation of USA, the current COVID-19 epicenter. The USA coronavirus response coordinator asked to take mental preparation for counting the death toll between the range of 100,00 to 200,000 in USA only from the prediction of best computer models, even after maintain the current strict social distancing (40). Koczkodaj et al., (2020) predicted about 10000000 COVID-19 cases outside of China by March 30, 2020 following the simple heuristic prediction method. The prediction of these researchers will be close to half of our prediction including China if it continues to April 29, 2020, the end date until which we continued our prediction.

The COVID-19 transmission shows a different trend of spread between the European temperate and Asian tropical countries. Though the first incident time has a gap of about one month, however the scenario between the countries of these two climatic regions has a head and tail difference. European countries is suffocating from the current COVID-19, a uncontrollable situation given applying modern medical facilities for their citizens by ensuring 11.5 critical care beds per 100,000 head of population (42) comparing with the only 3.6 critical care beds per 100,000 population in the entire Asian countries. The problem for European countries specifically in Italy and Spain is that they are getting 8.5 and 8.9 confirmed COVID-19 cases against per critical care bed (43), which is bounding them to take impossible choice of setting age limit for access to critical care bed. Though the European countries followed the same lockdown, test-trace-isolate infected ones’, social distance strategy to defeat COVID-19 like other Asian counterparts, however it was a bit delay to take initiatives by the decision-making authorities for slow, hesitant and in some cases even chaotic management being of their liberal democratic culture. Experts now also raised question about the effectuated tests quantity as well as quality variation between the two parts of the world when facing the same problem.

Moreover, there is criticism from the expert community for the individualistic hedonists of the Europeans, that dared the Government authorities to impose social distance or self-quarantine, which was quite easy in Asian countries for community-driven, and hierarchical nature of political and administrative intuitions. Not like war, the real war against pandemic COVID-19 the Europe’s 27 centers of decision and 27 lines of command also lambaste by their own parliament member (44).

Concerning to climatic pattern, there is also vice-versa prediction from the scientist and epidemiologist communities. Expert opinion is that the Asian countries is taking advantage from onset spring and will be favored on door knocking summer from COVID-19 cold shock for their geographical position and climatic condition. Lowen and Stell (45) argued that the droplet-mediated viral disease (SARS and MARS) survived in low absolute humidity (cold and dry weather) and attenuated the viral transmission in high absolute humidity (warm and humid weather), which can also be true for their genetic cousin COVID-19. However, Luo et al., (46) burst a vigilant caution that, the increasing temperature and humidity on the onset summer cannot hold back the COVID-19 transmission race too early. If anyhow the transmission is settled down in temperate countries taking the favor of high temperature and humidity, then Neher et al., (13) warned for having the chance of larger peak in the winter of 2020/21. Moreover, given the condition of very low spreads and fatality in the tropical countries, tend to be developing countries having very poor capacity of diagnosis and medical services, if the COVID-19 infections remain asymptotic until coming winter and then it respreads, it will be a toughest job in the planet for the millions of people to survive. Therefore, with the global leadership of the WHO, a comprehensive diagnosis of all the people irrespective of nationalities has to be implemented when going to get a window of opportunity for better preparation of health care systems, they must make hay while the sun shines by next six months. This will provide the information base for the individual countries to take the actions from the viewpoints of political action, social movement and health services.

## Conclusions and recommendations

Based on the analysis of secondary data in a predictive model, it was found that the that COVID- 19 spread and fatality tend to be high globally but compared to tropical countries, it is going to be incredibly high in the temperate countries for having lower temperature (7-16°C) and humidity (80-90%). However, some literature predicted that this might not be true, rather irrespective of weather conditions there might be a large number of asymptotic COVID-19 carrier in everywhere in the world particularly in the developing countries which have limited capacity of diagnosis and medical service, will be unknown unless the entire population is diagnosed. The asymptotic career might be the cause of outbreak in the coming winter while whether will be again favorable for COVID-19 incubation in both tropics of the world. Therefore, a comprehensive program with the leadership of WHO for testing of entire population of the world is required, which will be very useful for the individual government of member states of the United Nations to take proper political action and social and medical service. Specific and effective actions as recommended by WHO, UNICEF, CDC and ISGlobal (47–50) could be implemented to minimize the predicted COVID-19 moves are as follows.

- Eschew to touch different parts (eye, nose, mouth) of your face which facilitate the SARS-CoV-2 be wedged into your body especially its’ favorite lower part of the lungs. Also maintain personal hygiene at your best level by frequent washing your hands.
- Practice to regular use of disposable tissue when sneezing or coughing being COVID-19 a droplet-mediated viral disease and dispose it by maintaining proper disposing manner.
- Avoid the mass gathering and unpopular movement except for buying food and medicines. And when moving outside keep a 1 meter (3 feet) distance, especially when someone next to you is coughing or sneezing and vice-versa.
- The air born nature of the SARS-CoV-2 can be hindered by using consistent and correct use of surgical masks – must disinfect your hands after removing masks and gloves.
- Daily measure own temperature. Pursue medical attention through advance call when got fever, cough or difficult breathing. Therefore, the regional health care authority can provide the right health facility by protecting both the infected ones and other from spreading the virus.
- Install some kind of physical barrier (for example, Plexiglas shield) between the hospital reception staffs and patients. Also make adequate (1 meter) distance between the chairs in the waiting room of the emergency hospital visitors.
- Before making a home-call patient appointment ask do they have a cold, flu symptoms, fever, cough or conjunctivitis. And also ask about the recent contact with people suffering from COVID-19 infection or comes from infected areas. If so, recommend to wait for the visit or to go to the hospital (outpatients service, not emergency room).
- All units of hospitals and patient carrying ambulance will be cleaned regularly by using sterilized products as the virus has transmitted nature from the infected surface.
- And the last but not least, a simple message for all countries from the speech of WHO Director General “test, test, test”. This is for understating the scale of outbreak and reduce the transmission by following the strategy of test-trace-isolate infected ones.

All predictions, suggestions and efforts made by the researchers and health care professionals in different part of the worlds will frail without the implementation of extensive public health interventions.

## Data Availability

It was required two different types of data. Since for the prediction of daily new cases and deaths from COVID-19 the data from the official website of WHO (https://www.who.int/emergencies/diseases/novel-coronavirus-2019) and Center for Systems Science and Engineering, Johns Hopkins University, USA (https://www.arcgis.com/apps/opsdashboard/index.html#/bda7594740fd40299423467b48e9ecf6) were used. The weather data of temperate and tropical countries were gathered from authentic website of Weather Atlas (https://www.weather-atlas.com).

